# Medically relevant tandem repeats in nanopore sequencing of control cohorts

**DOI:** 10.1101/2024.03.06.24303700

**Authors:** Wouter De Coster, Ida Höijer, Inge Bruggeman, Svenn D'Hert, Malin Melin, Adam Ameur, Rosa Rademakers

## Abstract

Research and diagnostics for medically relevant tandem repeats and repeat expansions are hampered by the lack of population-scale databases. We attempt to fill this gap using our pathSTR web tool, which leverages long-read sequencing of large cohorts to determine repeat length and sequence composition in the general population. The current version includes 878 individuals of the 1000 Genomes Project cohort sequenced on the Oxford Nanopore Technologies PromethION. A comprehensive set of medically relevant tandem repeats were genotyped using STRdust to determine the tandem repeat length and sequence composition. PathSTR provides rich visualizations of this dataset, as well as the feature to upload one’s own data for comparison along the control cohort. We demonstrate the implementation of this application using data from targeted nanopore sequencing of a patient with Myotonic Dystrophy type 1. This resource will empower the genetics community to get a more complete overview of normal variation in tandem repeat length and sequence composition, and enable a better assessment of the pathogenic impact of tandem repeats observed in patients. PathSTR is available at https://pathstr.bioinf.be

## Introduction

Tandem repeats are defined as head-to-tail direct repetitions of a DNA motif, which can be repeated exactly, with motif interruptions or with an entirely different sequence composition for some alleles. Early methods involved (repeat-primed) PCR followed by Sanger sequencing or fragment length analysis and Southern blotting, which are low throughput, labor-intensive, and do not fully describe all repeat properties. Short-read sequencing technologies have difficulty in correctly determining their size, especially as the repeat length gets longer than the read length, but some specialized methods have enabled population-wide tandem repeat genotyping (Dashnow et al., 2021; Dolzhenko et al., 2019; Jam et al., 2023), allowing imputation and the identification of tandem repeats relevant for traits and diseases (Manigbas et al., 2024). Increased resolution, however, is offered by long-read sequencing technologies such as nanopore and PacBio sequencing, which enables direct observation of the repeat’s length, sequence composition, and DNA methylation. The field has yet to fully mature, and multiple genotyping, benchmarking, and comparison tools have been developed recently (Dolzhenko et al., 2023; English et al., 2023; Jam et al., 2024).

In the past years, these technologies have led to multiple novel discoveries of repeats associated with human disease (Cortese et al., 2023; Rafehi et al., 2023; Tan et al., 2023). To date, 68 repeats are associated with human diseases and summarized in STRchive (Dashnow, 2023/2023), although not all of these are firmly established as pathogenic. This is the set of repeats that we consider ‘medically relevant’ in the remainder of this manuscript. However, we anticipate this is only the tip of the iceberg of pathogenic tandem repeats. Both the repeat length and its composition are crucial determinants of the pathogenic potential of a specific repeat allele. A database of tandem repeat genotypes is beneficial to accurately assess an expanded allele’s pathogenic potential versus a common population polymorphism. Recently, long-read sequencing technologies have matured sufficiently to apply to population-scale sequencing projects (Beyter et al., 2021; De Coster et al., 2021; Noyvert et al., 2023). The genomics community has a long-standing tradition of making data freely available, which greatly benefits the interpretation of variants identified in patients, especially in the context of rare diseases. In this work, we describe pathSTR, a web app for the visualization of repeat length and sequence composition of medically relevant tandem repeats, also equipped with options to compare genotypes from other individuals (e.g. patients of interest) with the control cohort. At the time of writing, the database consists of genotypes of 878 individuals from the 1000 genomes project (1000 Genomes Project Consortium et al., 2015), sequenced on the Oxford Nanopore Technologies PromethION (Noyvert et al., 2023). The repeats are genotyped using STRdust, an efficient tandem repeat variant caller for long-read sequencing data. We provide this dataset to the genomics community for the improved interpretation of tandem repeat alleles.

## Results

### PathSTR visualization of repeat length and sequence composition

STRdust was used for genotyping the medically relevant tandem repeats from CRAM files over FTP using the curl feature of htslib(Bonfield et al., 2021). While an in-depth benchmark is outside of the scope of this work, we compared STRdust with TRGT(Dolzhenko et al., 2023) and LongTR(Jam et al., 2024) using the HG002 TR benchmark (English et al., 2023), demonstrating excellent concordance for tandem repeat lengths for the repeats of interest (Supplementary Fig. S1).

The pathSTR web app (https://pathstr.bioinf.be) shows the variation in tandem repeat length (Fig.1) and sequence composition (Fig. 2) across a large cohort of control individuals. Length plots, showing either the total length of the repeat or the difference with the reference genome, can be changed to show the pathogenic length cut-off (as obtained from STRchive), perform a log-transformation of the repeat length, show a density plot and split the individuals per 1000 Genomes super population and/or sex. The composition is visualized in three ways, either ‘raw’, showing the frequency of each motif, or ‘collapsed’, grouping samples with similar motif frequencies or ‘sequence’, showing the per-individual sequence of frequently-observed motifs across the repeat length. The repeat genotyping required only two reads to support an allele to maximize sensitivity, including at limited sequencing depth. Detailed information is available for further evaluation for each individual genotype in the web app, as well as the number of supporting reads and an IGV visualization of the alignment.

**Figure 1:**
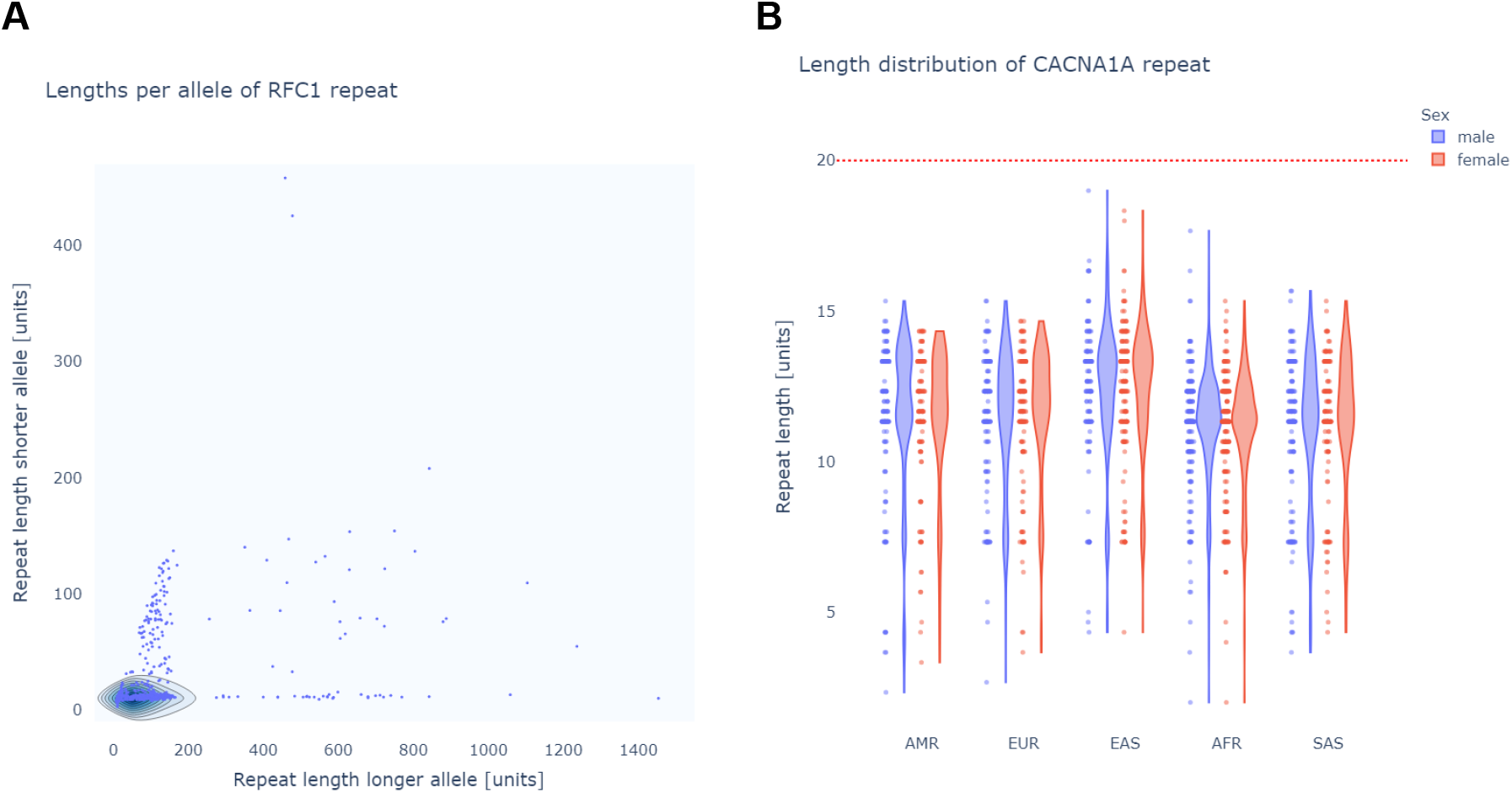
pathSTR visualization of the repeat length **A:** scatter plot length visualization of the *RFC1* repeat, comparing, per individual, the longer allele against the shorter. **B:** violin plots showing the *CACNA1A* repeat length, split by the 1000 Genomes Project super population (AMR: Admixed Americans, EUR: Europeans, EAS: East Asians, AFR: Africans, SAS: South Asians) and sex, showing the pathogenic repeat length from STRchive with a red horizontal line.

**Figure 2:**
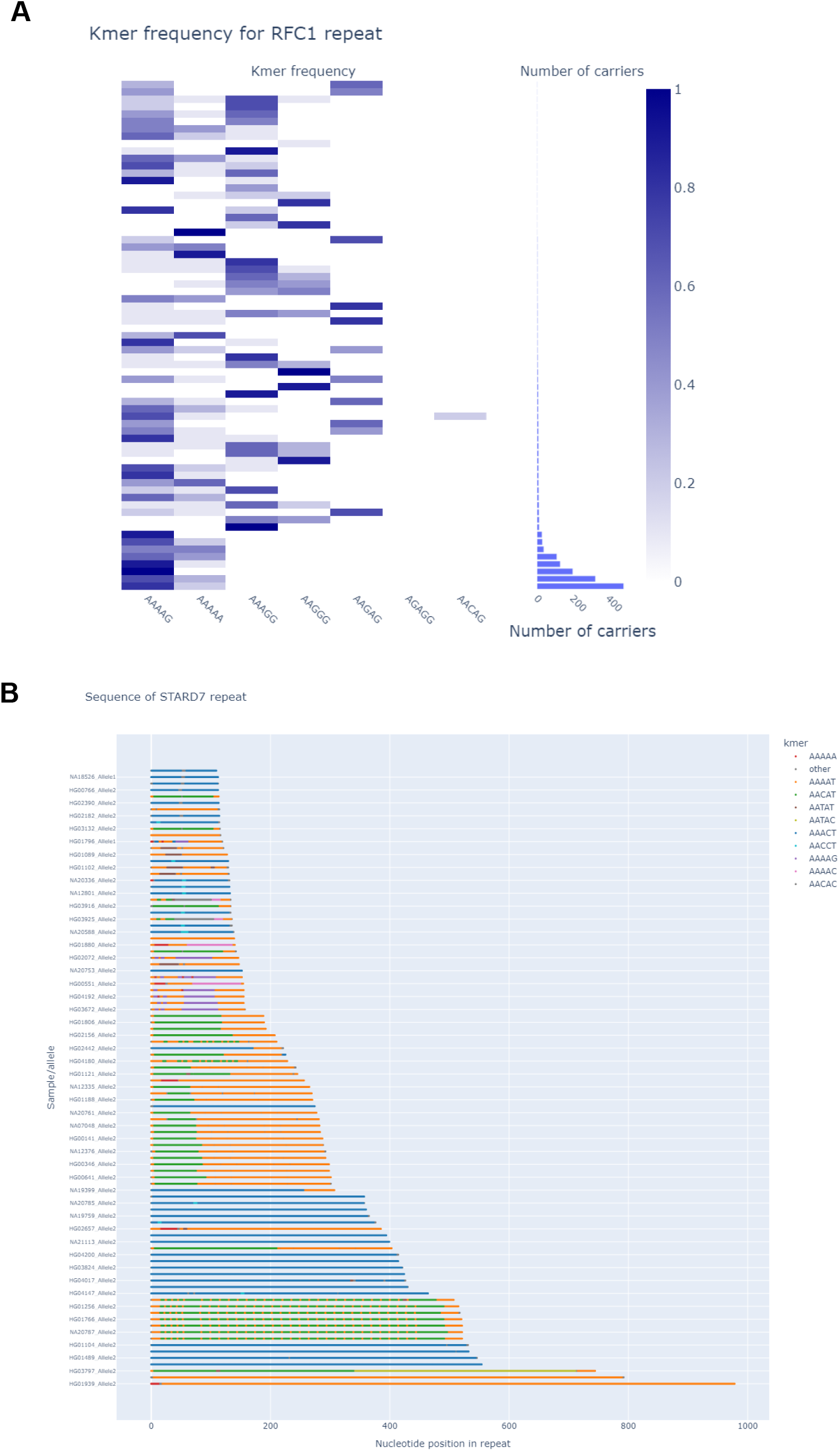
pathSTR visualization of the repeat sequence composition **A:** *RFC1* composition *collapsed* by motif, showing a marginal histogram to show the size of the groups, while hiding every combination of motifs that is only seen in a single individual. **B:** *sequence* plot of the *STARD7* repeat, showing colors for the most frequently seen motifs and grey for everything else, sorted by length. This example excludes the longest and shortest alleles.

### PathSTR to evaluate pathogenic repeats

STRchive includes some tandem repeats for which there is conflicting evidence for the association of this repeat expansion with a disease. One of these is a TTC repeat expansion in *DMD* linked to Duchenne Muscular Dystrophy, where it was suggested that >60 repeat units is pathogenic (Kekou et al., 2016). Investigation of this repeat using pathSTR (Fig. 3A) clearly supports the notion that the frequency of expanded alleles is too high to be causally linked to an early-onset condition, suggesting it is not a sufficient factor to explain the disease in the family in which the expansion was described.

**Figure 3:**
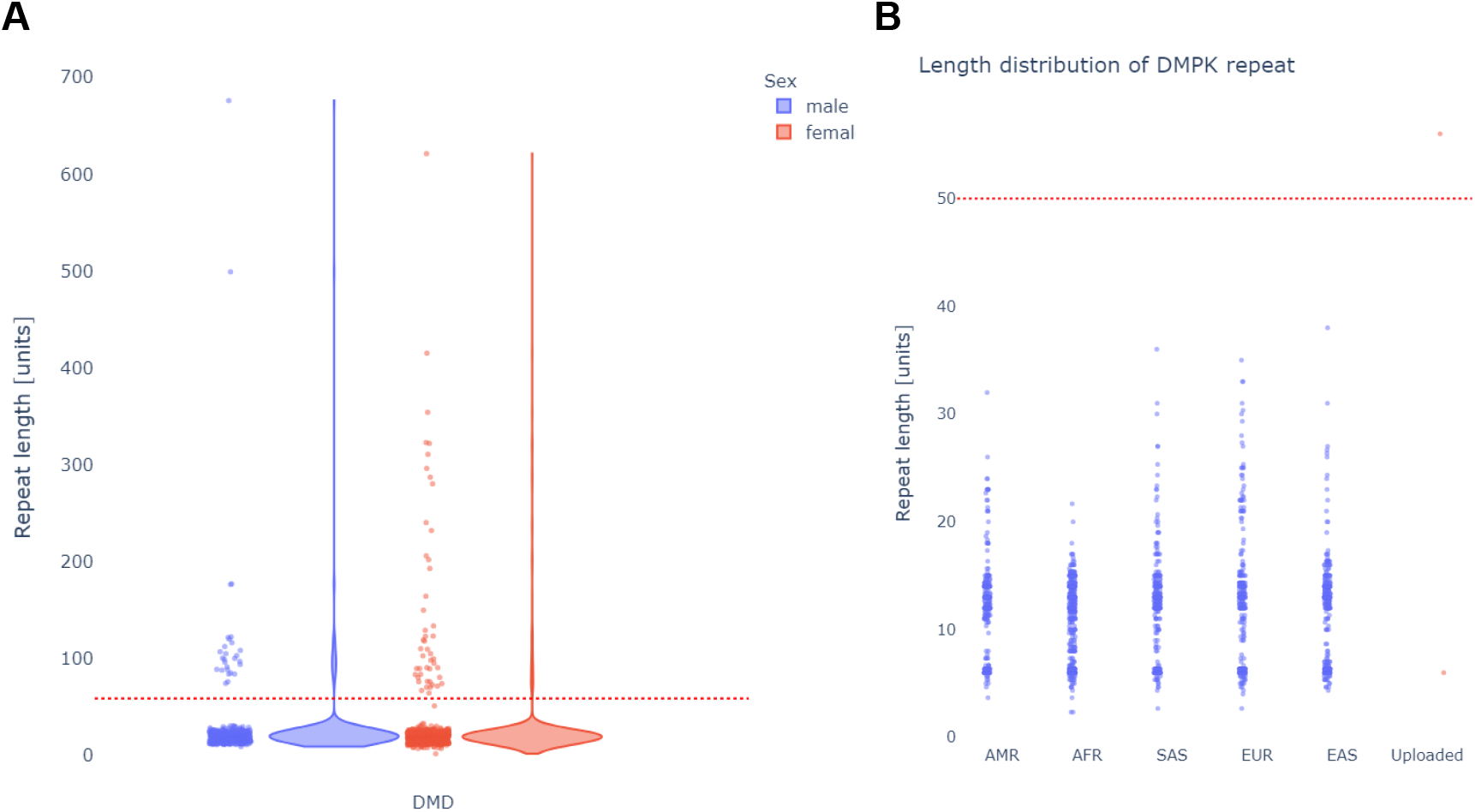
**A:** Repeat length in the *DMD* gene shows that many individuals in the general population have lengths above the proposed pathogenic length (red dotted line), suggesting it is not pathogenic. **B:** length visualization of the *DMPK* repeat, showing the uploaded data of the DM1 patient obtained using Cas9-enrichment, with the pathogenic cut-off indicated with a red dotted line.

PathSTR enables users to upload tandem repeat genotypes generated using STRdust from their own datasets, e.g. patients of interest, and will display those next to the control cohort. Fig. 3B shows an example of comparing Cas9-targeted resequencing data (see methods) (Gilpatrick et al., 2020) for a patient with Myotonic Dystrophy type 1 (DM1) with a DMPK expansion. PathSTR shows that the DMPK repeat length in this patient is indeed above the pathogenic line and is a length outlier compared to the rest of the cohort (z-score=9.5).

## Discussion

We present the data of medically relevant tandem repeats generated with publicly available long-read sequencing data from a control population, in a highly informative web application with rich visualizations of tandem repeat length and sequence composition. We prioritized sensitivity to detect long alleles for the pathSTR database, as the sequencing data of the 1000 genomes project used in this work were not uniformly sequenced to a high depth. We have demonstrated that STRdust is on par with other recently developed tandem repeat genotypers. However, an independent and more rigorous evaluation in this developing field is required. A first step towards this goal was already taken by compounding a tandem repeat catalog, assessing the tandem repeat variation for the HG002 Genome in a Bottle sample, and developing tools for comparison of methods (English et al., 2023).

While pathSTR should already greatly facilitate the interpretation of tandem repeats, we must stress that expert knowledge and caution are still required for the correct interpretation of results. The pathogenic length, as obtained from STRchive, must be evaluated in light of the available data, and may have to be reviewed when larger groups of patients and controls are sequenced. An additional reason for caution is that other repetitive sequences often flank tandem repeats, as is the case for *HTT*, where both repeats are often genotyped simultaneously (Höijer et al., 2018). This then complicates the assessment of the pathogenic character, as only the CAG/polyQ repeat is known to expand and cause disease. The flanking CCG/polyP repeat is stable but could confuse the genotyper, while simultaneously also influencing the polyQ pathogenicity (Urbanek et al., 2020). For these reasons, pathSTR will display a warning when the pathogenic length is added to the plots.

Sequence composition is another important determinant of pathogenicity. A clear example includes the intronic pentamer repeats in the *YEATS2* gene, (Supplementary Fig. S5), one of the causes of Familial adult myoclonic epilepsy (FAME), where only expanded repeats with an ATTTC motif are pathogenic. In this instance, evaluating patients on the overall repeat length alone is insufficient as expansions of ATTTC are pathogenic, but expansions of the reference sequence ATTTT are seen in healthy individuals (Depienne & Mandel, 2021). We are likely still lacking the full picture of the sequence compositions of these tandem repeats as long-read methods have only recently started probing the composition of expanded alleles. A more complete view of the sequence composition in expanded repeats of reference individuals and clinical cases will improve our understanding of what makes repeats pathogenic, and eventually lead to better diagnostics.

The current dataset does not provide information on the DNA methylation status, which can be determined from nanopore sequencing as native DNA is sequenced without amplification(Giesselmann et al., 2019). This would be a very relevant layer of information to incorporate in a later update, as especially long CG-rich repeats are known to be methylated and lead to epigenetic silencing in cis (Depienne & Mandel, 2021). This resource will continuously be expanded when new population sequencing efforts are made available online, as well as when additional tandem repeats are identified as relevant for human diseases.

## Methods

### Quality control

We used cramino for quality control and to determine library metrics such as library N50, yield, and normalized coverage per chromosome (De Coster & Rademakers, 2022). Samples with an estimated coverage <10x (32Gb) were removed (N=129, Supplementary Fig. S2), as well as a sample with unexpected normalized coverage on the sex chromosomes (Supplementary Figure S3).

### Genotyping medically relevant tandem repeats

Tandem repeats for pathSTR were genotyped with STRdust, which is implemented in Rust and uses the rust-htslib and rust-bio crates (Köster, 2016). STRdust is implemented in such a way that alignment files (in CRAM or BAM format) do not have to be available locally but can instead be queried from a remote location (using FTP, HTTPS, or s3), which is relevant in the context of this application. Alignments to an artificial reference sequence without the repeat sequence are done using the rust bindings to minimap2 (Guhlin, 2022/2024; Li, 2021), after which a consensus of the repeat allele is generated using a Partial Overlap Alignment (SPOA) (Vaser et al., 2017), as implemented in rust-bio. STRdust will perform pairwise alignment and hierarchical clustering to identify the reads that make up the two alleles and assign a heterozygous or homozygous genotype, excluding any read with a repeat sequence that does not result in an appropriate alignment with sequences from any of the other reads (outlier). STRdust does not attempt to split reads into two haplotypes for haploid sex chromosomes in male individuals. Commandline arguments are parsed with clap(*Clap-Rs/Clap: A Full Featured*, *Fast Command Line Argument Parser for Rust*, n.d.), and parallelization is achieved using rayon(*Rayon-Rs/Rayon*, 2014/2024). Binaries for STRdust are available at https://github.com/wdecoster/STRdust, with the source code available under the MIT license.

The repeats with a role in human diseases selected for genotyping were taken from STRchive (Dashnow, 2023/2023), using the hg38 coordinates for genotyping, the motif length for kmer composition plots, and the provided cut-off for repeats to be considered pathogenic. STRdust implements a --pathogenic option to download tandem repeat coordinates from STRchive for ease of genotyping these medically relevant tandem repeats. Genotyping the 1000 Genomes samples is organized using the snakemake workflow manager (Koster & Rahmann, 2012).

We compared STRdust (v0.5.0) to TRGT (v0.8.0-11fecab) and LongTR (version 638942f-dirty), for the set of medically relevant tandem repeats and evaluated the correlation of the obtained repeat allele lengths with a scatter plot matrix and calculating the Parson correlation.

### PathSTR web app

The pathSTR web app is written in Python and built upon dash (*Dash Documentation & User Guide* | *Plotly*, n.d.) and additionally uses cyvcf2 to parse VCF files (Pedersen & Quinlan, 2017), pandas to manipulate data frames (McKinney, 2011), and modules from the Python standard library (*Python Programming Language* | *USENIX*, n.d.). The parsed data is saved into an hdf5 container for easier access and quick start-up times (The HDF Group, 2020/2024). For every repeat and individual, an IGV visualization is provided using igv.js(Robinson et al., 2023) as made available through dash-bio (Hossain, 2019). The pathSTR web app is hosted in-house, deployed using nginx and gunicorn.

The repeat composition is analyzed by counting kmers in the repeat sequence according to the forward strand of the reference genome, splitting sequences based on the known repeat motif length but unbiased to which motifs can be found. Each kmer is rotated (GCA-CAG-AGC) and represented by either the known unit (as defined by STRchive) or the alphabetically first motif of the rotations. Motifs that are seen too rarely (with cutoffs depending on the visualization type) are discarded, as we assume those are sequencing noise. The ‘sequence’ visualization will still show the unassigned rare motifs in grey for inspection (Supplementary Fig. S4). The code of the pathSTR app is open source and available under the MIT license at https://github.com/wdecoster/pathSTR

### Targeted nanopore sequencing

5 μg of genomic DNA was extracted from whole blood for Cas9 targeted sequencing using the SQK-CS9109 kit (Oxford Nanopore Technologies) according to the manufacturer’s protocol. Nanopore sequencing was performed on a MinION R9.4.1 flowcell (Oxford Nanopore Technologies) at SciLifeLab National Genomics Infrastructure (NGI) in Uppsala, Sweden. Basecalling was done with Guppy (ONT), followed by alignment to the hg38 reference genome with minimap2.

## Data Availability

The data generated in this project can be accessed at https://pathstr.bioinf.be, where the data can be queried, visualized, and downloaded in the form of a tab-separated file or individual VCF files as generated by STRdust. The original sequencing data is available at https://ftp.1000genomes.ebi.ac.uk/vol1/ftp/data_collections/1KG_ONT_VIENNA/hg38/

## Ethics declaration

The study concerning the DM1 patient was approved by the Swedish Ethical Review Authority (2019-04746), and written informed consent was obtained from each participating individual or their respective legal guardians.

## Competing interest statement

WDC has received free consumables and travel reimbursement from Oxford Nanopore Technologies.

## Acknowledgments

We thank Noyvert and co-authors for making their data freely available. WDC is a recipient of a postdoctoral fellowship from FWO [12ASR24N]. WDC thanks the staff from the NGI Uppsala Genome Center for the warm welcome and pleasant research stay.

## Notes

### Author Declarations

The study concerning the DM1 patient was approved by the Swedish Ethical Review Authority (2019-04746), and written informed consent was obtained from the participating individual or their respective legal guardians.

## References

1000 Genomes Project Consortium, Auton, A., Brooks, L. D., Durbin, R. M., Garrison, E. P., Kang, H. M., Korbel, J. O., Marchini, J. L., McCarthy, S., McVean, G. A., & Abecasis, G. R. (2015). A global reference for human genetic variation. Nature, 526(7571), 68–74. 10.1038/nature15393

Beyter, D., Ingimundardottir, H., Oddsson, A., Eggertsson, H. P., Bjornsson, E., Jonsson, H., Atlason, B. A., Kristmundsdottir, S., Mehringer, S., Hardarson, M. T., Gudjonsson, S. A., Magnusdottir, D. N., Jonasdottir, A., Jonasdottir, A., Kristjansson, R. P., Sverrisson, S. T., Holley, G., Palsson, G., Stefansson, O. A., … Stefansson, K. (2021). Long-read sequencing of 3,622 Icelanders provides insight into the role of structural variants in human diseases and other traits. Nat. Genet., 53(6), 779–786. 10.1038/s41588-021-00865-4

Bonfield, J. K., Marshall, J., Danecek, P., Li, H., Ohan, V., Whitwham, A., Keane, T., & Davies, R. M. (2021). HTSlib: C library for reading/writing high-throughput sequencing data. Gigascience, 10(2). 10.1093/gigascience/giab007clap-rs/clap: A full featured, fast Command Line Argument Parser for Rust. (n.d.). Retrieved February 13, 2024, from https://github.com/clap-rs/clap

Cortese, A., Beecroft, S. J., Facchini, S., Curro, R., Cabrera-Serrano, M., Stevanovski, I., Chintalaphani, S., Gamaarachchi, H., Weisburd, B., Folland, C., Monahan, G., Scriba, C. K., Dofash, L., Johari, M., Grosz, B. R., Ellis, M., Fearnley, L. G., Tankard, R., Read, J., … Ravenscroft, G. (2023). A CCG expansion in ABCD3 causes oculopharyngodistal myopathy in individuals of European ancestry (p.2023.10.09.23296582). medRxiv. 10.1101/2023.10.09.23296582

Dash Documentation & User Guide | Plotly. (n.d.). Retrieved February 13, 2024, from https://dash.plotly.com/

Dashnow, H. (2023). Hdashnow/STRchive [HTML]. https://github.com/hdashnow/STRchive (Original work published 2023)

Dashnow, H., Pedersen, B. S., Hiatt, L., Brown, J., Beecroft, S. J., Ravenscroft, G., LaCroix, A. J., Lamont, P., Roxburgh, R. H., Rodrigues, M. J., Davis, M., Mefford, H. C., Laing, N. G., & Quinlan, A. R. (2021). STRling: A k-mer counting approach that detects short tandem repeat expansions at known and novel loci. In bioRxiv. 10.1101/2021.11.18.469113

De Coster, W., & Rademakers, R. (2022). NanoPack2: Population scale evaluation of long-read sequencing data. In bioRxiv. 10.1101/2022.11.28.518232

De Coster, W., Weissensteiner, M. H., & Sedlazeck, F. J. (2021). Towards population-scale long-read sequencing. Nat. Rev. Genet. 10.1038/s41576-021-00367-3

Depienne, C., & Mandel, J.-L. (2021). 30 years of repeat expansion disorders: What have we learned and what are the remaining challenges? Am. J. Hum. Genet. 10.1016/j.ajhg.2021.03.011

Dolzhenko, E., Bennett, M. F., Richmond, P. A., Trost, B., Chen, S., van Vugt, J. J. F. A., Nguyen, C., Narzisi, G., Gainullin, V. G., Gross, A., Lajoie, B., Taft, R. J., Wasserman, W. W., Scherer, S. W., Veldink, J. H., Bentley, D. R., Yuen, R. K. C., Bahlo, M., & Eberle, M. A. (2019). ExpansionHunter Denovo: A computational method for locating known and novel repeat expansions in short-read sequencing data. In bioRxiv. 10.1101/863035

Dolzhenko, E., English, A., Dashnow, H., Brandine, G. D. S., Mokveld, T., Rowell, W. J., Karniski, C., Kronenberg, Z., Danzi, M. C., Cheung, W. A., Bi, C., Farrow, E., Wenger, A., Martínez-Cerdeño, V., Bartley, T. D., Jin, P., Nelson, D., Zuchner, S., Pastinen, T., … Eberle, M. A. (2023). Resolving the unsolved: Comprehensive assessment of tandem repeats at scale (p. 2023.05.12.540470). bioRxiv. 10.1101/2023.05.12.540470

English, A., Dolzhenko, E., Jam, H. Z., Mckenzie, S., Olson, N. D., Coster, W. D., Park, J., Gu, B., Wagner, J., Eberle, M. A., Gymrek, M., Chaisson, M. J. P., Zook, J. M., & Sedlazeck, F. J. (2023). Benchmarking of small and large variants across tandem repeats (p. 2023.10.29.564632). bioRxiv. 10.1101/2023.10.29.564632

Giesselmann, P., Brändl, B., Raimondeau, E., Bowen, R., Rohrandt, C., Tandon, R., Kretzmer, H., Assum, G., Galonska, C., Siebert, R., Ammerpohl, O., Heron, A., Schneider, S. A., Ladewig, J., Koch, P., Schuldt, B. M., Graham, J. E., Meissner, A., & Müller, F.-J. (2019). Analysis of short tandem repeat expansions and their methylation state with nanopore sequencing. Nat. Biotechnol., 1–4. 10.1038/s41587-019-0293-x

Gilpatrick, T., Lee, I., Graham, J. E., Raimondeau, E., Bowen, R., Heron, A., Downs, B., Sukmar, S., Sedlazeck, F. J., & Timp, W. (2020). Targeted nanopore sequencing with Cas9-guided adaptor ligation. Nature Biotechnology, 38(4), 433–438. 10.1038/s41587-020-0407-5

Guhlin, J. (2024). Jguhlin/minimap2-rs [Rust]. https://github.com/jguhlin/minimap2-rs (Original work published 2022)

Höijer, I., Tsai, Y.-C., Clark, T. A., Kotturi, P., Dahl, N., Stattin, E.-L., Bondeson, M.-L., Feuk, L., Gyllensten, U., & Ameur, A. (2018). Detailed analysis of HTT repeat elements in human blood using targeted amplification-free long-read sequencing. Human Mutation, 39(9), 1262–1272. 10.1002/humu.23580

Hossain, S. (2019). Visualization of Bioinformatics Data with Dash Bio. Proceedings of the 18th Python in Science Conference, 126–133. 10.25080/Majora-7ddc1dd1-012

Jam, H. Z., Li, Y., DeVito, R., Mousavi, N., Ma, N., Lujumba, I., Adam, Y., Maksimov, M., Huang, B., Dolzhenko, E., Qiu, Y., Kakembo, F. E., Joseph, H., Onyido, B., Adeyemi, J., Bakhtiari, M., Park, J., Javadzadeh, S., Jjingo, D., … Gymrek, M. (2023). A deep population reference panel of tandem repeat variation. In bioRxiv. 10.1101/2023.03.09.531600

Jam, H. Z., Zook, J. M., Javadzadeh, S., Park, J., Sehgal, A., & Gymrek, M. (2024). Genome-wide profiling of genetic variation at tandem repeat from long reads (p. 2024.01.20.576266). bioRxiv. 10.1101/2024.01.20.576266

Kekou, K., Sofocleous, C., Papadimas, G., Petichakis, D., Svingou, M., Pons, R.-M., Vorgia, P., Gika, A., Kitsiou-Tzeli, S., & Kanavakis, E. (2016). A dynamic trinucleotide repeat (TNR) expansion in the DMD gene. Molecular and Cellular Probes, 30(4), 254–260. 10.1016/j.mcp.2016.07.001

Koster, J., & Rahmann, S. (2012). Snakemake—A scalable bioinformatics workflow engine. Bioinformatics, 28(19), 2520–2522. 10.1093/bioinformatics/bts480

Li, H. (2021). New strategies to improve minimap2 alignment accuracy. In arXiv [q-bio.GN]. http://arxiv.org/abs/2108.03515

Manigbas, C., Jadhav, B., Garg, P., Shadrina, M., Lee, W., Martin-Trujillo, A., & Sharp, A. (2024). A phenome-wide association study of tandem repeat variation in 168,554 individuals from the UK Biobank (p. 2024.01.22.24301630). medRxiv. 10.1101/2024.01.22.24301630

McKinney, W. (2011). pandas: A foundational Python library for data analysis and statistics. Python for High Performance and Scientific Computing, 1–9.

Noyvert, B., Erzurumluoglu, A. M., Drichel, D., Omland, S., Andlauer, T. F. M., Mueller, S., Sennels, L., Becker, C., Kantorovich, A., Bartholdy, B. A., Braenne, I., Bolivar-Lopez, J. C., Mistrellides, C., Belbin, G. M., Li, J. H., Pickrell, J. K., Jong, J. de, Arora, J., Hu, Y., … Ding, Z. (2023). Imputation of structural variants using a multi-ancestry long-read sequencing panel enables identification of disease associations (p. 2023.12.20.23300308). medRxiv. 10.1101/2023.12.20.23300308

Pedersen, B. S., & Quinlan, A. R. (2017). cyvcf2: Fast, flexible variant analysis with Python. Bioinformatics, 33(12), 1867–1869. 10.1093/bioinformatics/btx057 Python Programming Language | USENIX. (n.d.). Retrieved February 13, 2024, from https://www.usenix.org/conference/2007-usenix-annual-technical-conference/presentation/python-programming-language

Rafehi, H., Read, J., Szmulewicz, D. J., Davies, K. C., Snell, P., Fearnley, L. G., Scott, L., Thomsen, M., Gillies, G., Pope, K., Bennett, M. F., Munro, J. E., Ngo, K. J., Chen, L., Wallis, M. J., Butler, E. G., Kumar, K. R., Wu, K. H., Tomlinson, S. E., … Lockhart, P. J. (2023). An intronic GAA repeat expansion in FGF14 causes the autosomal-dominant adult-onset ataxia SCA27B/ATX-FGF14. The American Journal of Human Genetics, 110(1), 105–119. 10.1016/j.ajhg.2022.11.015

Rayon-rs/rayon. (2024). [Rust]. rayon-rs. https://github.com/rayon-rs/rayon (Original work published 2014)

Robinson, J. T., Thorvaldsdottir, H., Turner, D., & Mesirov, J. P. (2023). igv.js: An embeddable JavaScript implementation of the Integrative Genomics Viewer (IGV). Bioinformatics, 39(1), btac830. 10.1093/bioinformatics/btac830

Tan, D., Wei, C., Chen, Z., Huang, Y., Deng, J., Li, J., Liu, Y., Bao, X., Xu, J., Hu, Z., Wang, S., Fan, Y., Jiang, Y., Wu, Y., Wu, Y., Wang, S., Liu, P., Zhang, Y., Yang, Z., … Xiong, H. (2023). CAG Repeat Expansion in THAP11 Is Associated with a Novel Spinocerebellar Ataxia. Movement Disorders, 38(7), 1282–1293. 10.1002/mds.29412

The HDF Group. (2024). Hierarchical Data Format, version 5 [C]. https://github.com/HDFGroup/hdf5 (Original work published 2020)

Urbanek, A., Popovic, M., Morató, A., Estaña, A., Elena-Real, C. A., Mier, P., Fournet, A., Allemand, F., Delbecq, S., Andrade-Navarro, M. A., Cortés, J., Sibille, N., & Bernadó, P. (2020). Flanking Regions Determine the Structure of the Poly-Glutamine in Huntingtin through Mechanisms Common among Glutamine-Rich Human Proteins. Structure (London, England: 1993), 28(7), 733–746.e5. 10.1016/j.str.2020.04.008

Vaser, R., Sović, I., Nagarajan, N., & Šikić, M. (2017). Fast and accurate de novo genome assembly from long uncorrected reads. Genome Res., 27(5), 737–746. 10.1101/gr.214270.116

