## Supplementary data for "Medically relevant tandem repeats in nanopore sequencing of control cohorts"

### Supplementary Figures

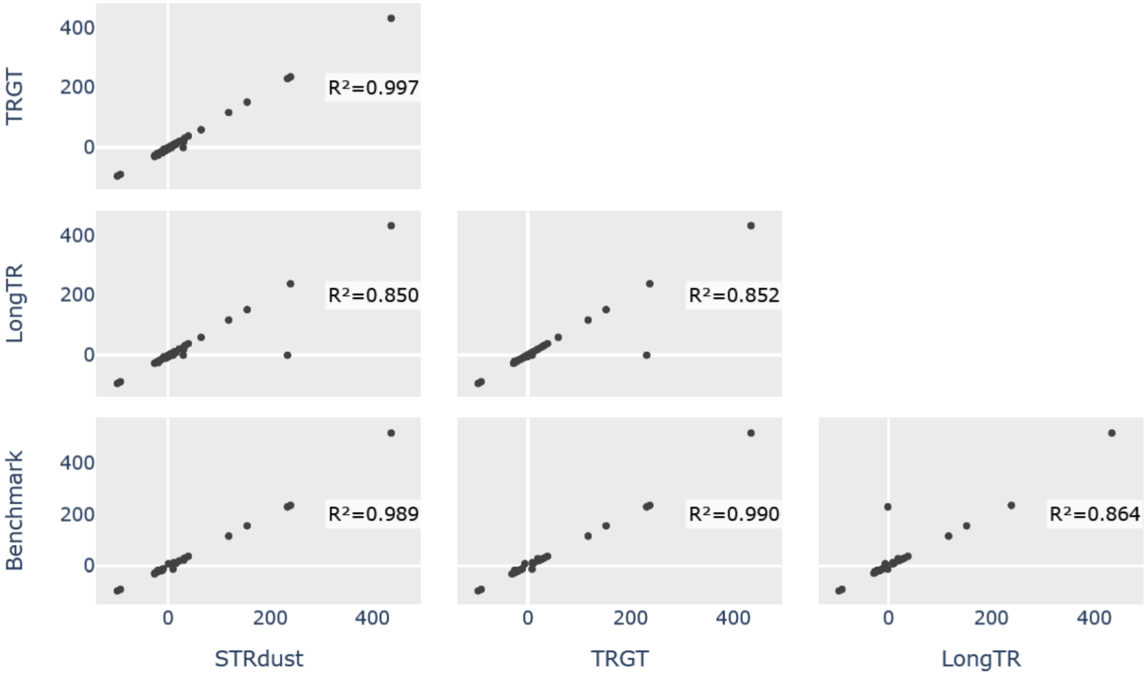


**Supplementary Figure S1**: correlation of medically relevant tandem repeat lengths across genotypers, compared to the HG002 benchmark lengths.

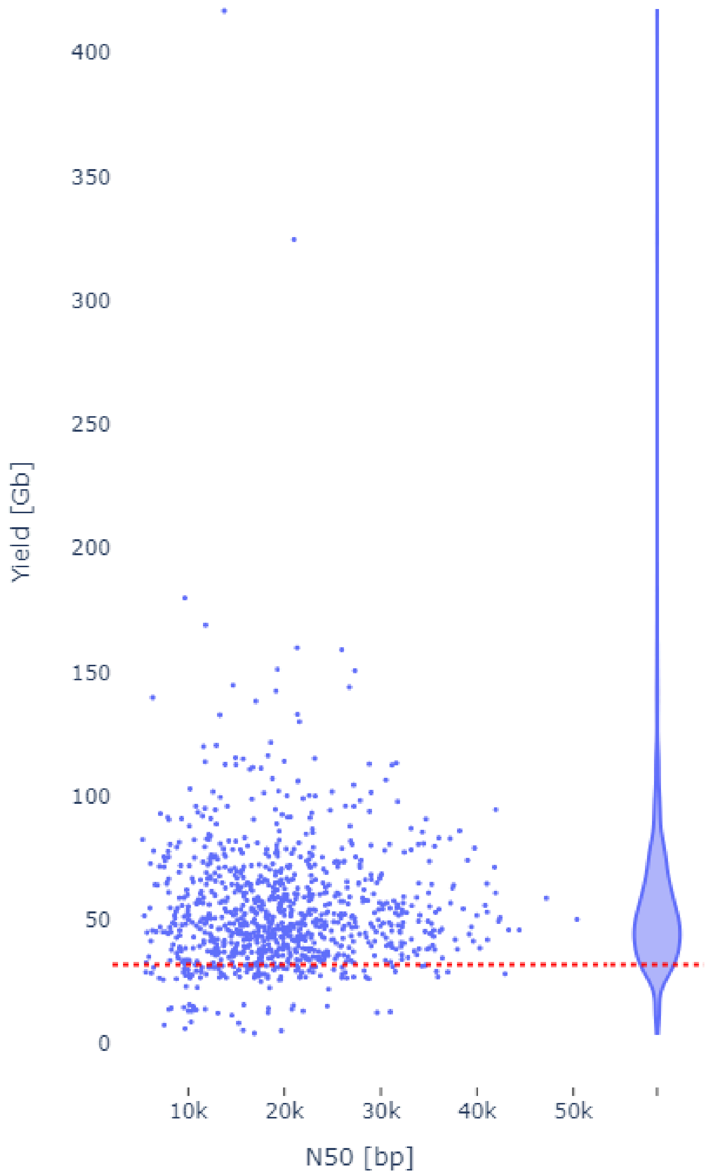


Supplementary Figure S2: scatter plot of sequencing yield and N50 library length, with a marginal violin plot for yield and a horizontal line for the minimum yield cutoff (32Gb).

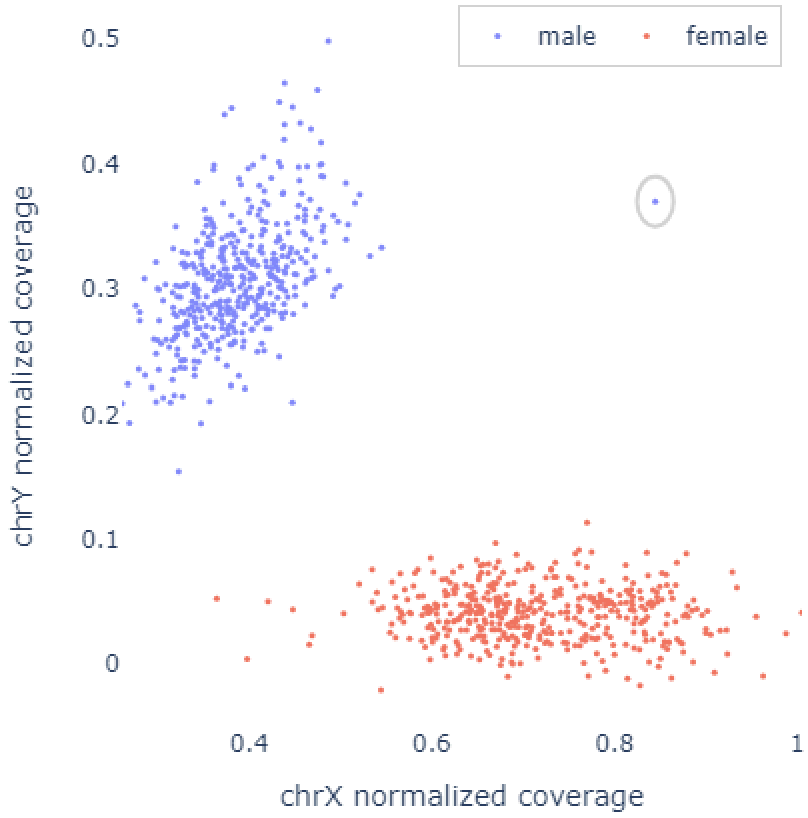


**Supplementary Figure S3**: visualization of sex chromosome dosage, with a circle around one suspected carrier of Klinefelter syndrome (XXY, HG02372). Points are colored based on the sex as provided in the sample info file downloaded from https://www.internationalgenome.org/data-portal/sample

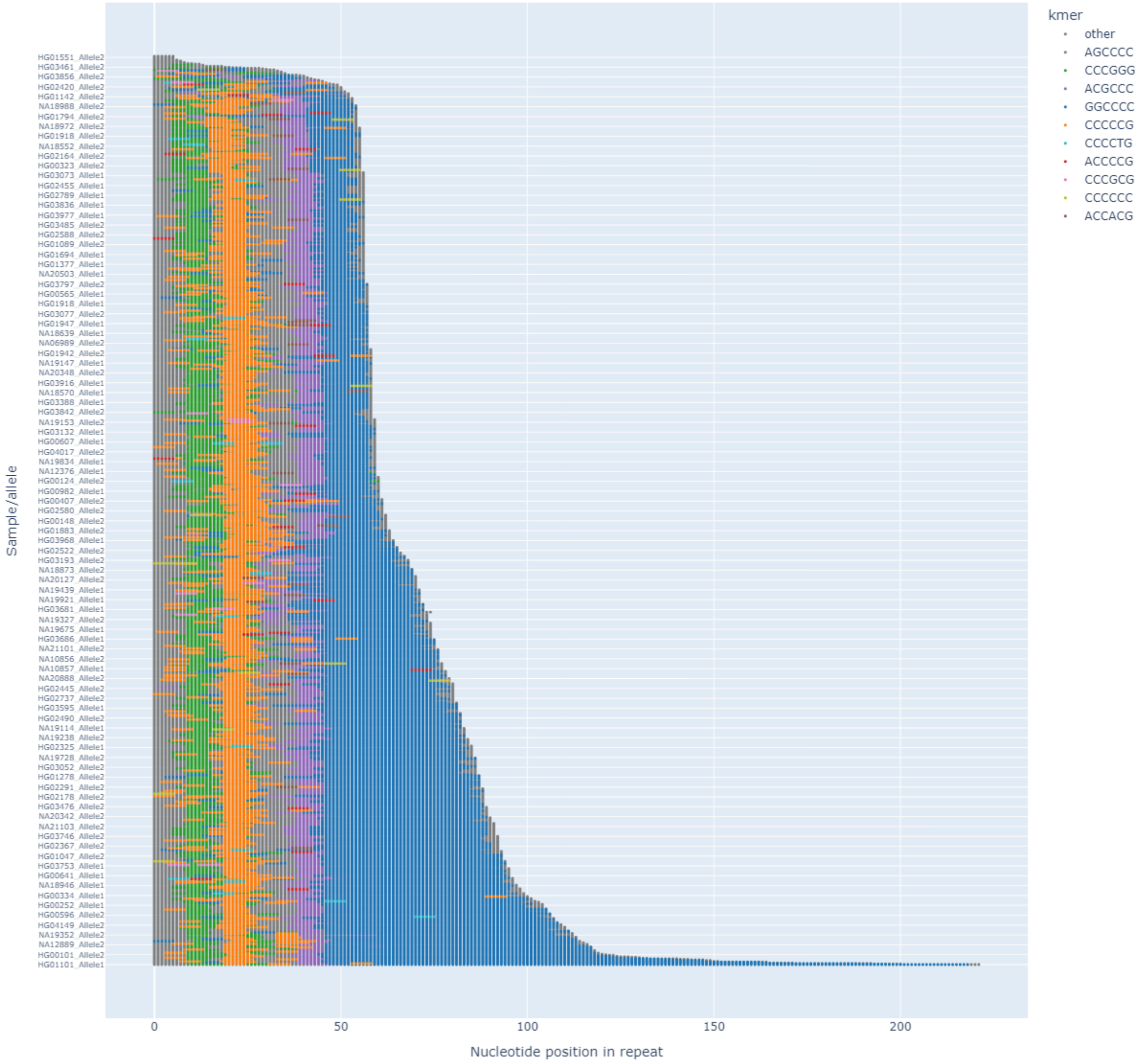


**Supplementary Figure S4**: *sequence* plot of the *C9orf72* repeat, showing colors for the most frequently seen motifs and grey for everything else, sorted by length.

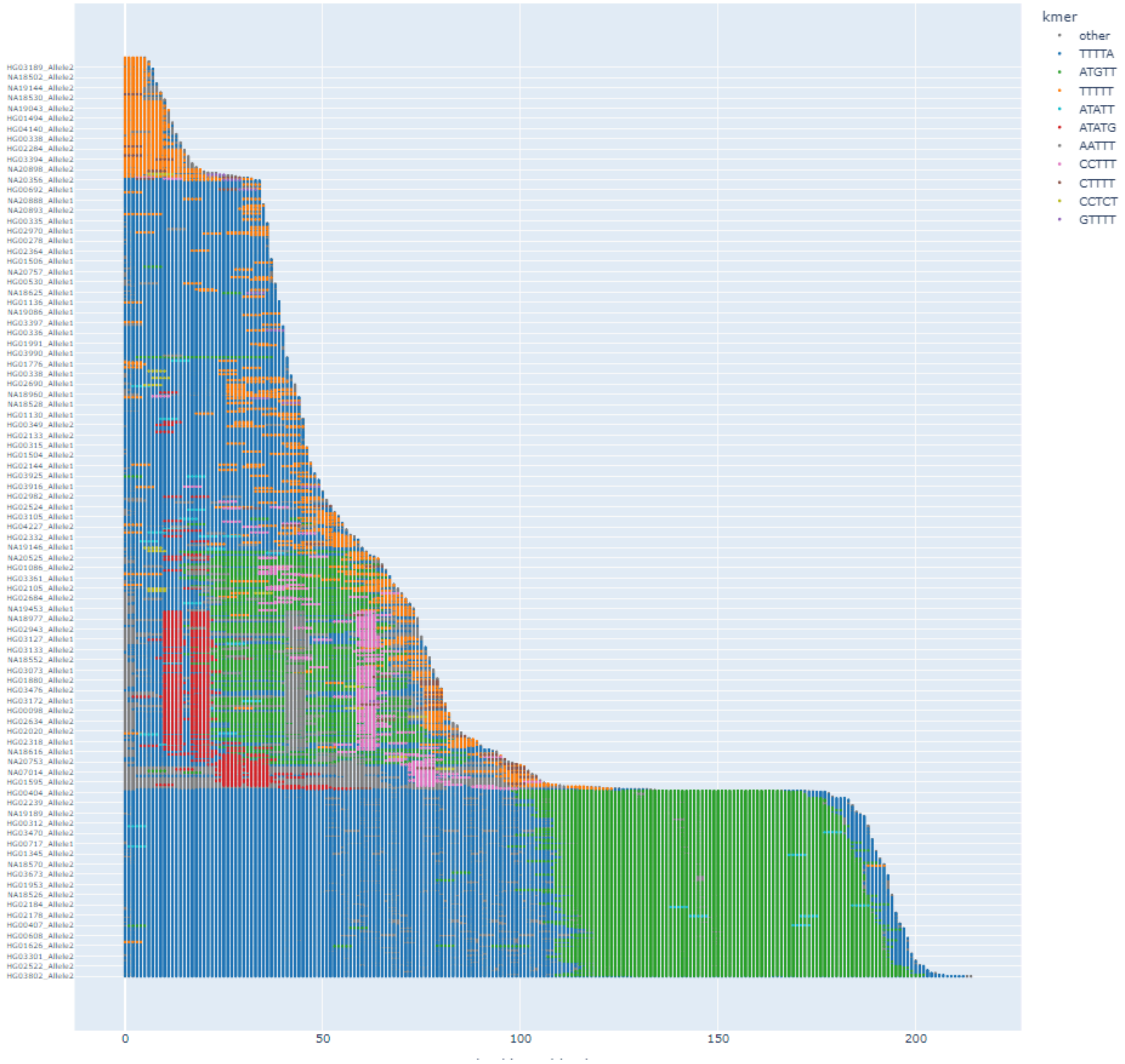


**Supplementary Figure S5**: *sequence* plot of the *YEATS2* repeat, showing colors for the most frequently seen motifs and grey for everything else, sorted by length.
